# Cardiorespiratory signature of neonatal sepsis: Development and validation of prediction models in 3 NICUs

**DOI:** 10.1101/2022.09.28.22280469

**Authors:** Sherry L. Kausch, Jackson G. Brandberg, Jiaxing J. Qiu, Aneesha Panda, Alexandra Binai, Joseph Isler, Rakesh Sahni, Zachary A. Vesoulis, J. Randall Moorman, Karen D. Fairchild, Douglas E. Lake, Brynne A. Sullivan

**Author notes:** Corresponding Author: Brynne A. Sullivan, PO Box 800386, Charlottesville, VA 22908.

## Abstract

**Background:** Heart rate characteristics aid early detection of late-onset sepsis (LOS), but respiratory data contain additional signatures of illness due to infection. Predictive models using cardiorespiratory data may improve early sepsis detection. We hypothesized that heart rate (HR) and oxygenation (SpO_2_) data contain signatures that improve sepsis risk prediction over HR or demographics alone.

**Methods:** We analyzed cardiorespiratory data from very low birth weight (VLBW, <1500g) infants admitted to three NICUs. We developed and externally validated four machine learning models to predict LOS using features calculated every 10m: mean, standard deviation, skewness, kurtosis of HR and SpO_2_, and cross-correlation. We compared feature importance, discrimination, calibration, and dynamic prediction across models and cohorts. We built models of demographics and HR or SpO_2_ features alone for comparison with HR-SpO2 models.

**Results:** Performance, feature importance, and calibration were similar among modeling methods. All models had favorable external validation performance. The HR-SpO_2_ model performed better than models using either HR or SpO_2_ alone. Demographics improved the discrimination of all physiologic data models but dampened dynamic performance.

**Conclusions:** Cardiorespiratory signatures detect LOS in VLBW infants at 3 NICUs. Demographics risk-stratify, but predictive modeling with both HR and SpO_2_ features provides the best dynamic risk prediction.

## Introduction

Early detection of late-onset sepsis (LOS, sepsis beyond 3 days of age) reduces mortality and improves outcomes for survivors ^1,2^. Although many infants with LOS exhibit clinical instability, signs and symptoms are nonspecific and occur too late in the course of illness. Signatures of illness are present in physiologic time series data derived from heart rate (HR) and oxygen saturation (SpO_2_) monitoring in the early stages of sepsis in premature infants^3–5^. We developed and validated algorithms to detect abnormal patterns in continuous HR^4^ and SpO_2_^6^ data and in their cross-correlation ^3^.

The heart rate characteristics (HRC) index estimates the risk of imminent sepsis using the standard deviation of RR intervals, sample asymmetry^7^, and sample entropy^8^ to detect decreased HR variability with transient decelerations^4,9^. A multicenter randomized clinical trial showed HRC display reduced mortality for premature infants^1,10^.

We have found additional physiological signatures of neonatal illness and explored new data collection, analysis, and modeling methods^3,5,6^. Respiratory deterioration prompts many sepsis evaluations^5^ in premature infants, and apnea increases during sepsis^11^ as inflammation affects central control of breathing^12^. Thus, we hypothesize that abnormal patterns in pulse oximetry might add to HR characteristics in early detection of sepsis. Here, we used data from three tertiary Neonatal ICUs to develop and validate statistical models combining HR and SpO_2_ analytics for sepsis detection in very low birth weight (VLBW, <1500g) infants. We aimed to evaluate multiple cardiorespiratory features, modeling methods, and performance metrics to test the hypothesis that respiratory data contain signatures of illness caused by sepsis and add information to HR characteristics and demographic variables for risk prediction.

When we can determine that signatures of illness are present, we can contemplate using statistical models as bedside predictive tools. Here, we refer to models of this kind as POWS (Pulse Oximetry Warning System).

## Methods

### Patients

We studied VLBW infants admitted to 3 NICUs: University of Virginia Children’s Hospital (NICU 1, 2012-2021), Morgan Stanley Children’s Hospital of New York, Columbia University (NICU 2, 2012-2019), and St. Louis Children’s Hospital, Washington University School of Medicine (NICU 3, 2016-2021). We report the results of this study in accordance with the Transparent Reporting of a multivariable prediction model for Individual Prognosis Or Diagnosis (TRIPOD) guidelines^13,14^.

The Institutional Review Boards of each institution approved the study. We excluded infants with major chromosomal or congenital anomalies and those with no vital sign data collected. We collected demographic and clinical variables from the electronic health record and unit databases.

### Sepsis definition

Late-onset septicemia (LOS) is the primary outcome of our modeling. Clinicians at each site reviewed blood cultures obtained after three days of age for infants who met inclusion criteria. We recorded the event as LOS if the blood culture was positive, the infant was treated with at least five days of antibiotics, and the culture was preceded by at least two days with no antibiotics. We excluded negative blood cultures, positive blood cultures obtained within seven days of a prior positive blood culture, and positive blood cultures treated as contaminants (defined as <5 days of antibiotics).

### HR and SpO2 data collection and preprocessing

Continuous HR and SpO_2_ data were collected from standard NICU bedside monitors (GE, Philips) using the BedMaster system (Hillrom’s Medical Device Integration Solution, Chicago, IL; formerly Excel Medical, Jupiter, FL). Electrocardiogram-derived HR and pulse oximeter-derived SpO_2_ and pulse rate were collected at 0.5 Hz at NICU 1 and 2. NICU 3 collected data at 1 Hz and was downsampled to 0.5 Hz to match the other sites. SpO_2_ was measured with the default averaging time of 8 seconds, using Masimo technology at NICU 1 and 2 and Nellcor Oximax technology at NICU 3. HR and SpO_2_ underwent single-step preprocessing where values representing incontrovertible artifact (zeros) were removed.

### Candidate predictors: HR, SpO2, Demographics

In 10-minute non-overlapping windows, we calculated the mean, standard deviation (SD), skewness, kurtosis of both HR and SpO_2_, and their minimum and maximum cross-correlation ^3,5^. We examined the empirical univariate risk of each feature to evaluate individual predictor-risk relationships. We noticed differences in distributions of mean HR between the three sites. Additionally, prior work suggests mean HR does not predict imminent LOS^15^. Therefore, we excluded mean HR as a candidate predictor. Furthermore, including mean HR reduced performance across sites.

### Model development

Fig 1 provides a schematic overview of the methods, including model development, validation, comparisons, and secondary analyses. Every 10-minute window of raw data was labeled as “control,” “LOS,” or censored. Windows in the 24-hour period preceding the time of positive blood culture were labeled LOS, those falling in the seven days following LOS were censored, and all other windows falling between 72h after birth and NICU discharge were labeled as control. Windows with more than 50% data missing were excluded. All 10-minute windows labeled as LOS and a sub-sample of those labeled as control (one 10-minute window per hour) were used for training. We trained models using data from infants at NICU 1 and tested the models on NICU 2 and 3. Modeling was performed in R (R Foundation for Statistical Computing, Vienna, Austria) and Python (Python Software Foundation, https://www.python.org/).

**Fig 1.**
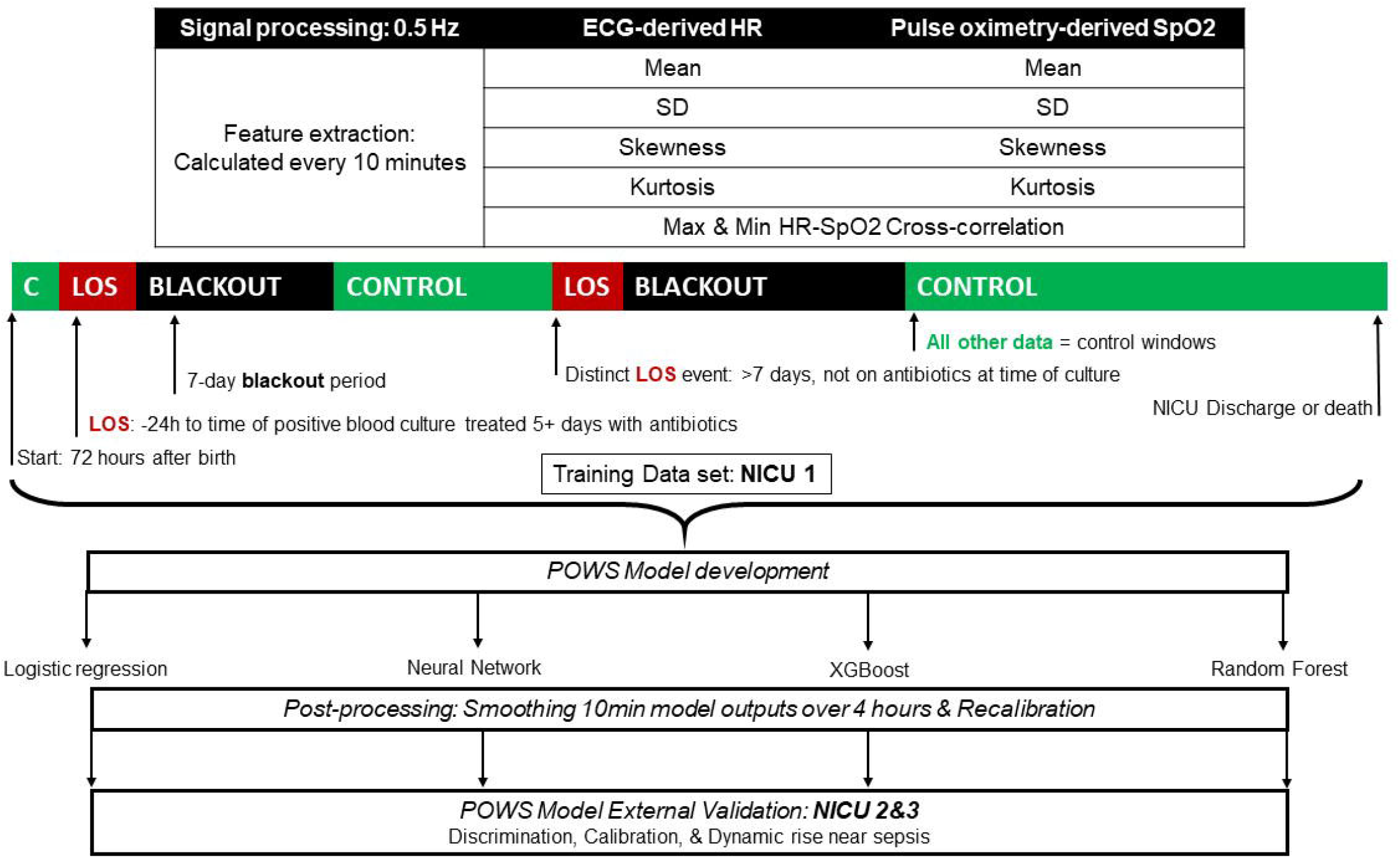
A Schematic Overview of Methods. From top to bottom, we processed the raw signals, sampled at 0.5 Hz, by calculating HR and SpO_2_ features every ten minutes. Each 10-minute window, from 72 hours after birth until NICU discharge or death, was labeled as late-onset sepsis (LOS), control, or removed as a blackout period window. Data from NICU 1 were used to train four machine learning models. Before external validation on data at NICU 2 & 3, post-processing steps included smoothing the 10-minute model outputs over 4 hours and recalibrating. Metrics used for external validation included discrimination by AUC, calibration, and plotting the average relative risk over the 48 hours preceding sepsis to look for a dynamic rise from baseline near the time of diagnosis by blood culture.

Each model generates the estimated probability of LOS, updated every 10-minutes. To translate model outputs into the fold-increased risk of LOS in the next 24 hours, we divided the estimated probability by the average probability of LOS. We evaluated different smoothing windows that averaged the 10-minute model outputs over different time periods, ranging from the preceding 1 to 12 hours. We determined an optimum soothing window by balancing the rise in area under the receiver operating characteristic (AUC) against the decrease in the average risk score for LOS. After smoothing, model outputs were sampled hourly. Finally, hourly, smoothed model predictions were recalibrated using the sigmoid function.

To determine importance, features were permuted to calculate the loss in AUC and ranked from highest to lowest. We added ranks across models to estimate the overall feature importance.

### Modeling methods

Four modeling strategies were evaluated, including logistic regression, a neural network, an ensemble method (extreme gradient boosting classifier or XGBoost), and random forest (Fig 1).

For model #1, we built an 18-dimensional logistic regression model by including one additional non-linear term^16,17^ for each of the nine features. We adjusted for repeated measures using the Huber-White method^18^. We removed minimum cross-correlation, SD SpO_2_, and non-linear effects with p-values >0.05. The result was an 11-dimensional model.

For model #2, we developed a neural network model with four hidden layers. These layers had 512, 256, 128, and 64 hidden neurons, respectively. Dropout layers at a rate of 0.3 were added as a regularization measure between each hidden layer ^19^. Leaky ReLU was used as the activation function in the hidden layers and a sigmoid activation function was used in the last layer ^20^. The model was trained using binary cross-entropy as the loss function and Adam as the optimizer with a learning rate of 0.001. An early stopping callback was used to avoid overfitting which monitored the loss with a patience of 10.

Model #3 was an Extreme Gradient Boosting (XGBoost) classifier ^21^. We trained multiple models on a parameter grid to identify optimal hyperparameters for learning rate, alpha (l1 regularization), lambda (l2 regularization), and max depth based on the highest validation AUC on NICU 2 after training on NICU 1. The model used binary logistic as its objective function.

Model #4 was a random forest model with 800 classification trees and the square root of the number of features were sampled as candidates at each split ^22^. The output of the model was the fraction of trees that classified an outcome as an event.

### Model validation

We validated models on unseen data from two independent sites. We trained models on NICU 1, the largest and most complete data set, and externally validated on data from the other sites. All subsequent analyses of model validation and performance used the models trained on data from NICU 1. We assessed model discrimination by AUC with confidence intervals based on 200 bootstrap runs.

We evaluated model calibration by plotting the predicted vs. observed relative risk of LOS. We assessed model performance in the critical period near the time of LOS diagnosis by examining the time course of the average relative risk. We identified a rise in risk by performing a sign-rank test with the null hypothesis that hourly risk estimates are equal to risk estimates from the same patient 24 hours prior with a statistical significance threshold of p<0.05.

### Sensitivity analyses

We assessed the dynamic change in the model output near the time of other “sepsis-like” events, including negative blood cultures diagnosed as clinical sepsis or necrotizing enterocolitis (NEC) without bacteremia. Because mechanical ventilation can alter HR and SpO_2_ patterns^3,23,24^, we also examined differences in models’ predicted probability of sepsis based on ventilator status. Given the potential confounding of coagulase-negative staphylococcus (CONS), ^25,26^ we conducted a subgroup analysis to assess model performance in CONS versus non-CONS positive blood cultures.

We tested whether models performed equivalently using features derived only from pulse-oximetry data. While ECG and pulse oximetry monitoring are standard, resource-poor settings might benefit from an algorithm that operates from an oximeter alone. Therefore, we tested model performance using features derived from pulse rate rather than the features derived from ECG HR.

### Model comparisons

We hypothesized that HR, SpO_2_, and static demographic variables are independent predictors of LOS. To quantify the added value, we built three additional models. First, we trained a demographics model that included birth weight, sex, and chronological age as baseline risk factors^4,15,27,28^. Second, we created a HR model that contained SD, skewness, and kurtosis. Third, we developed an SpO_2_ model that contained mean, SD, skewness, and kurtosis. For comparison to a validated measure, we also calculated the HRC index ^1,4^.

At a range of thresholds, we calculated the number of alerts per patient day, where an alert had to start within the day preceding the clinical diagnosis of sepsis and could only fire once per day. We excluded alerts in the seven days following a sepsis event.

## Results

### Participants

We studied 3,151 VLBW infants at the three sites. Vital sign data were available for 2494/3151 (79%) infants and 302 of 390 episodes of LOS (77%). Table 1 shows the characteristics of the study population. They are typical of a VLBW cohort, with minor inter-center differences. The median age of LOS was 16 days (IQR: 8 - 32). As expected, infants with LOS had lower gestational age and birth weight.

### Model development

We trained models on 923 infants from NICU 1. The nine candidate predictors and their empirical relationship with the outcome of sepsis are shown in S1 Fig. The retained features used in the logistic regression model were mean SpO_2_, SD HR, skewness of HR, skewness of SpO_2_, kurtosis of HR, kurtosis of SpO_2_, and maximum positive value of the HR-SpO_2_ cross-correlation. Table S2 presents model coefficients.

After models were trained, we engaged in three post-processing steps. First, we evaluated different smoothing windows that averaged the 10-minute model outputs over different time periods. The optimum soothing window balanced the rise in AUC against the decrease in the average risk score for LOS. The optimal smoothing window was four hours (S2 Fig). After smoothing all model outputs over 4 hours, we sampled the model outputs hourly (rather than every 10-minutes). Finally, we recalibrated prior outputs prior to performance evaluation and validation.

### Model performance

All modeling methods discriminated sepsis windows from control windows with good performance (AUCs > 0.8, Table 2), and external validation showed only small performance diminishment. Additionally, model performance was similar whether using ECG-derived HR or pulse rates from oximetry (AUC loss -0.001, -0.006, -0.013 at NICU 1, 2, and 3, respectively). There was high and consistent calibration (Fig 2). When the observed risk of sepsis is less than 1, all models slightly underpredicted risk at NICU 2 and overpredicted at NICU 3.

**Fig 2.**
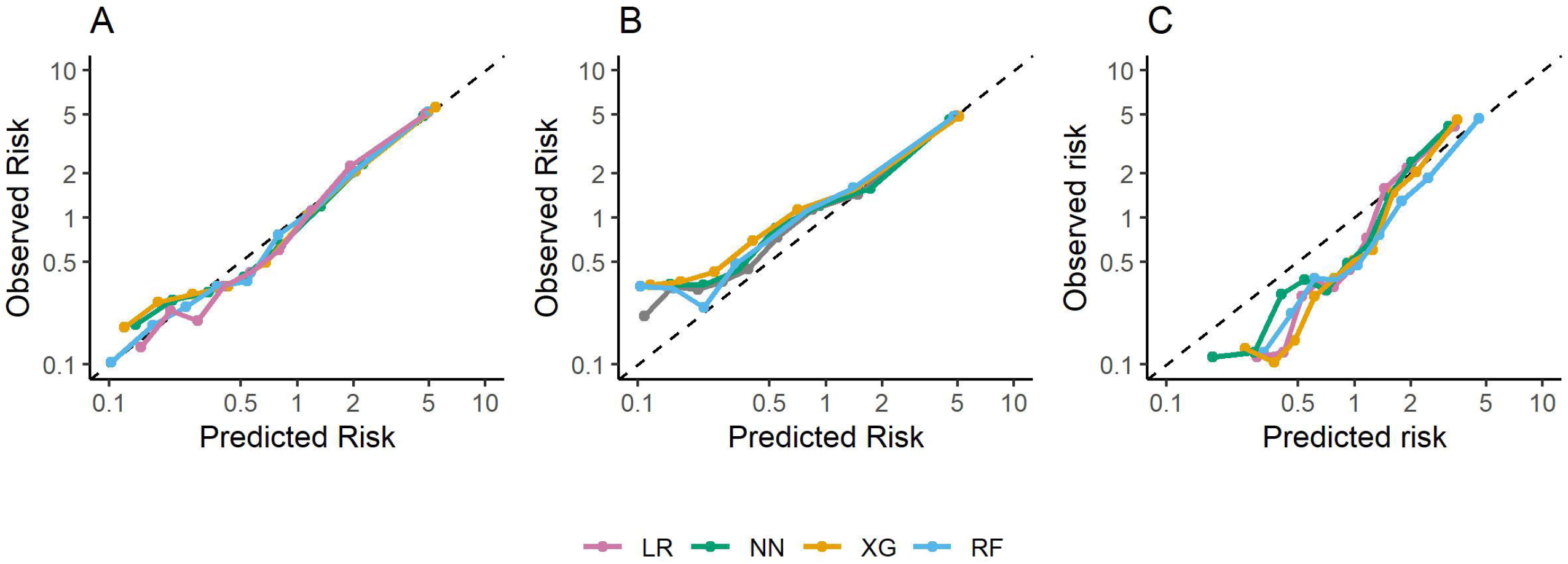
Calibration plots. Calibration of each POWS model for (A) NICU 1, (B) NICU 2, and (C) NICU 3. Model outputs are smoothed over 4 hours, sampled hourly, and recalibrated after smoothing. Predicted risk relative to average is on the abscissa and observed risk relative to average is on the ordinate. Each point represents one decile of predicted risk. The line of identity is shown as a dashed line. LR = logistic regression, NN = neural network, XG = XGBoost, RF = random forest

Fig 3 plots the average fold-increase in each risk model as a function of time to LOS at each NICU. A fold-increase of 1 indicates no greater risk for sepsis than the baseline (0.26%); a fold-increase of 2 would indicate twice the average daily risk (0.52%). The relative risk predicted by the logistic regression model increased by 92% in the 24 hours before sepsis (2.5-fold to 4.8-fold), and the predicted risk calculated in the XGBoost model increased by 150%, from 3.2-fold to 8.0-fold. Predicted risk values deviated significantly from baseline from 23 to 24 hours before the blood culture (Fig 3).

**Fig 3.**
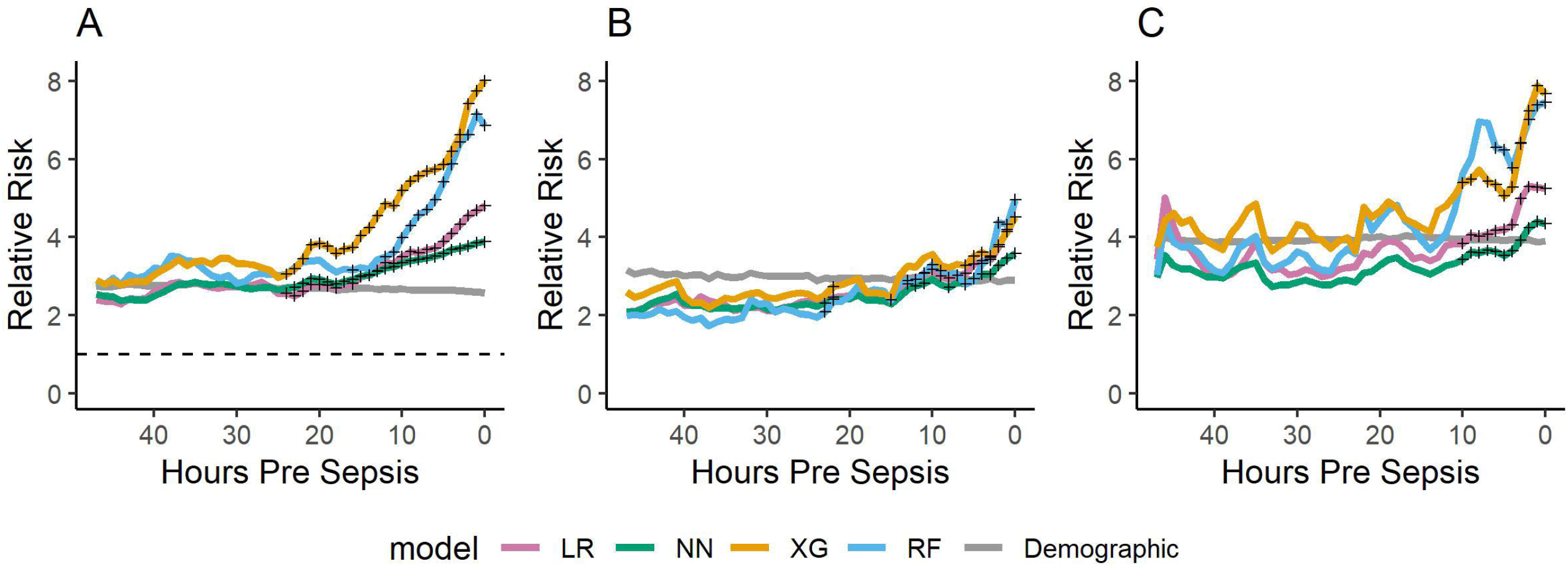
The Average Relative Risk of Sepsis. The average relative risk of sepsis as predicted by each model as a function of the time to event in hours. Panels show the results of each model at (A) NICU 1, (B) NICU 2, and (C) NICU 3. Results are shown for the four POWS models and the demographic-only model (in gray). Black crosses indicate times where the model outputs are significantly higher (plJ<lJ0.05) than outputs from the same patient 24lJh prior. LR = logistic regression, NN = neural network, XG = XGBoost, RF = random forest

### Feature importance

Fig 4 shows feature importance in the logistic regression (A), neural net (B), XGBoost (C), and random forest (D) sepsis prediction models. Features are ordered by decrease in AUC introduced by permuting each feature. Of note, seven features were included in the logistic regression model, while nine features were used in the remaining models. The features of greatest importance, identified by adding the ranks of the features across models, were skewness of HR, SD of HR, kurtosis of SpO_2_, and maximum cross-correlation. The random forest model identified kurtosis of SpO_2_ as the feature of greatest importance, while the remaining models ranked it as less important than skewness of HR and SD of HR.

**Fig 4.**
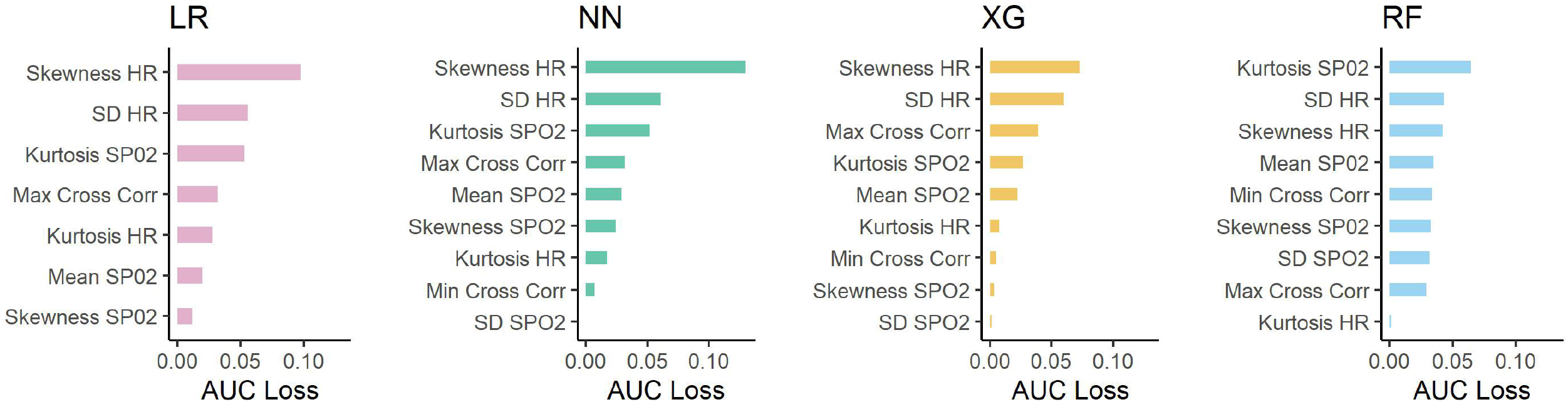
Variable importance plots. Variable importance plots for components of the logistic regression (LR), neural net (NN), XG Boost (XG), and random forest (RF) sepsis prediction models. Features are ordered by decreasing AUC loss introduced by permuting the values of each feature. Summing the rank of each feature across all four models gives the following overall rank to each feature: 1) Skewness HR, 2) SD HR, 3) Kurtosis SpO_2,_ 4) Max XC, 5) Mean SpO_2_, 6) Kurtosis HR, 7) Skewness SpO_2_, 8) Min XC, 9) SD SpO_2_.

### Do HR-SpO2 models add information to HR and demographics?

Having established that there is a cardiorespiratory signature of neonatal sepsis, we evaluated the potential clinical utility of the statistical models. We chose the logistic regression model, and we will call it POWS, for Pulse Oximetry Warning System.

We compared POWS to models with features derived from only HR data, only SpO_2_ data, or from only demographics of chronological age, sex, and birth weight. The parameters for each of these models are shown in supplementary tables.

Table 3 shows the results of model discrimination for LOS in training and testing data. The POWS model performed better than models using only HR-derived (AUC, 0.82 vs 0.80 NICU 1) or only SpO_2_-derived features (AUC, 0.82 vs 0.71, NICU 1) or only demographics (AUC 0.82 vs 0.78 NICU 1). As expected, the demographics model provides only near-static information (Figure 3).

To further understand how combining HR and SpO_2_ data features in prediction models adds to HR characteristics alone, we compared POWS to the HRC index, a logistic regression model that uses only features derived from HR data^4^. Although POWS and HRC both utilize HR-derived features, POWS uses every two-second HR data to calculate HR SD, skewness, and kurtosis, while the HRC index uses inter-beat-intervals to calculate HR SD, sample asymmetry, and sample entropy. Despite these differences, POWS performed similarly to the HRC index in the NICU 1 cohort (HRC index AUC 0.795, CI 0.798 - 0.803). Fig 5 displays predictiveness curves of the HRC index and POWS model. POWS fits the data better for prediction of LOS in the NICU 1 cohort.

**Fig 5.**
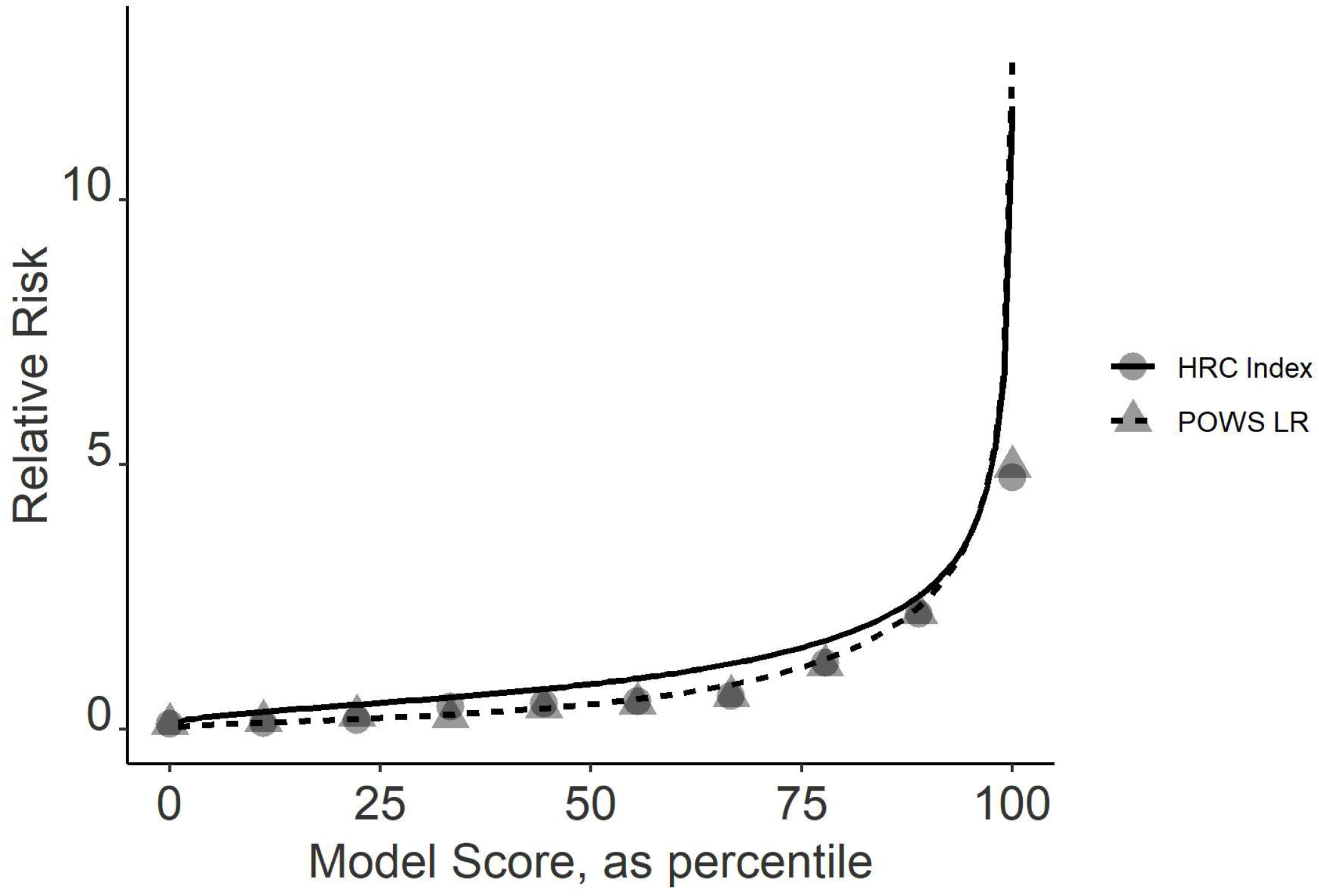
Predictiveness Curves. Predictiveness curves for HRC index score and POWS LR score in estimating sepsis risk. The solid lines show risk scores arrayed from smallest to largest. The circles are the observed fold in-increase in risk of sepsis. Blue represents the HRC index, and red represents the POWS LR scores.

### Sensitivity and alarm rates

Examining the sensitivity of the models across a range of thresholds, based on the number of daily alarms in a 50-bed unit, demonstrates that POWS has a perceptibly higher sensitivity than the models using HR or SpO2 features alone (Fig 6, Fig S5). The low sensitivity of the demographics model again speaks to the limitations of static variables for continuous risk prediction. Adding demographic features to POWS decreases the sensitivity across the range of alert rates and therefore limits its utility as an early warning score.

**Fig 6.**
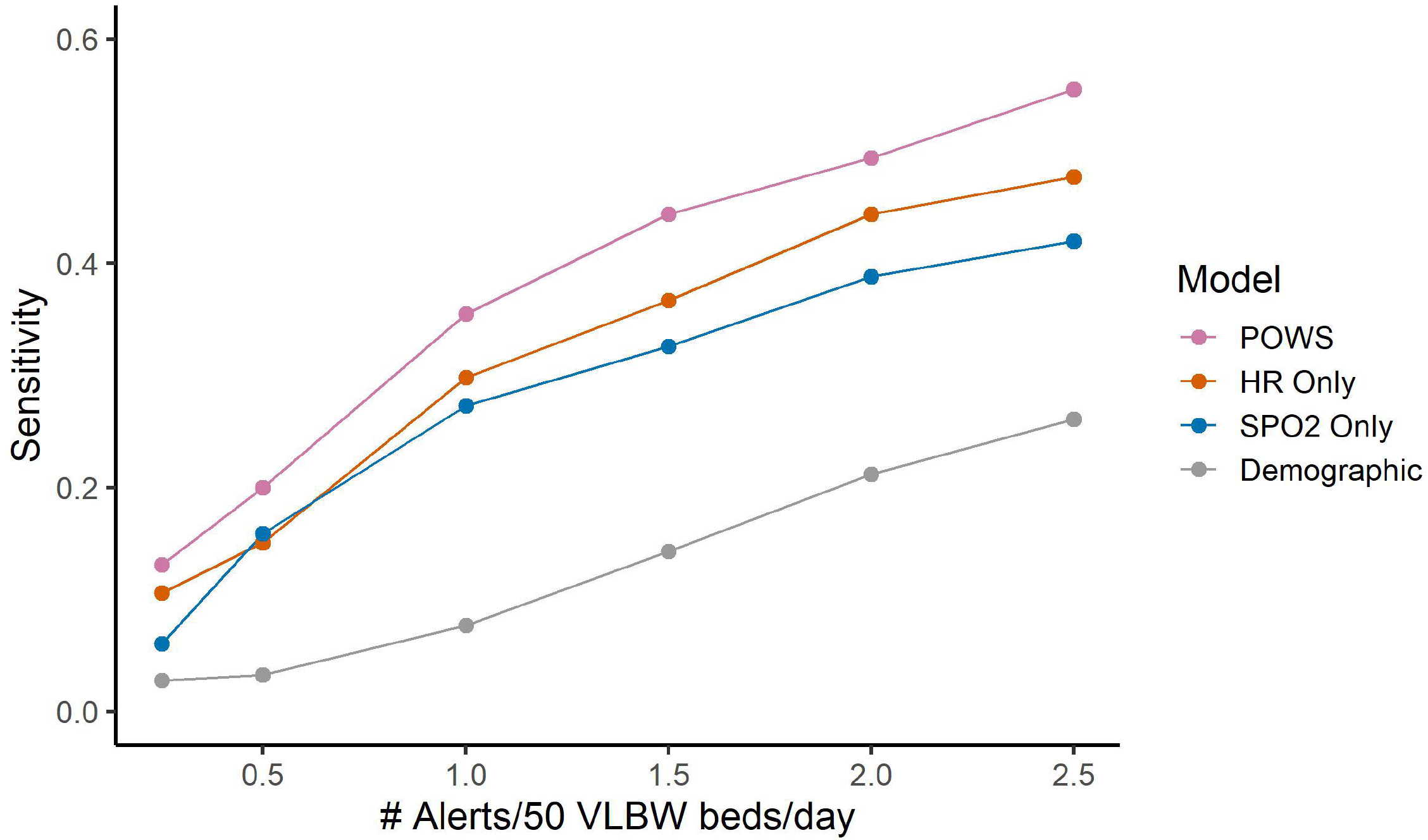
Evaluating the Sensitivity of Models across a Range of Thresholds. We selected a range of thresholds and then calculated the number of alerts per day and required an alert to start within the day preceding the clinical diagnosis of sepsis. We defined alerts as daily threshold crossings. We excluded alerts in the seven days following a sepsis event. The y-axis displays the percent of sepsis events detected when allowing for different numbers of alerts per day. The HRC index (not displayed) performs similarly to the HR-only model.

We examined lift charts as a way to visually compare the ability of each model to detect LOS events (Fig S6). Lift is the ratio of the percent of events captured at a given percentile of data to the random expectation of the percent of events captured. POWS had a higher lift than HR-only models, and both had a greater lift than the SpO_2_-only or demographics-only models. We performed this analysis to demonstrate the added value of combined cardiorespiratory modeling to detect sepsis within the highest risk segments of data.

### Model performance in other clinical contexts

We trained our models on blood-culture positive sepsis, but assessed model performance around the time of clinical deterioration with other diagnoses and stratified by factors that denote illness severity. First, we examined the dynamic risk estimates of POWS near the time of blood cultures for clinical sepsis (negative blood culture treated with antibiotics for at least five days due to clinical illness) and the time of blood cultures for confirmed NEC without bacteremia. Fig S3 shows a steep rise in risk in the hours preceding cultures for clinical sepsis and NEC, with a slower decrease in risk in the 48 hours following cultures.

Fig S4 shows model performance near LOS events, stratified by ventilator status at the time of sepsis and by causative organism, grouped as CONS vs. non-CONS. On average, LOS episodes on a ventilator and those caused by non-CONS bacteremia had higher risk scores before, during, and after diagnosis of LOS.

## Discussion

We show that cardiorespiratory signatures provide important information for early warning of LOS in VLBW infants. Prior work identified abnormal heart rate characteristics and increased HR-SpO_2_ cross-correlation as physiomarkers of illness due to sepsis ^3,5,29^. Here, we report that more information exists in the data at hand. Using machine learning modeling on multicenter data, we tested the hypothesis that HR and SpO_2_ data contain patterns that contribute independent, additive information for LOS detection. In doing so, we developed and validated cardiorespiratory models that predict an increased risk of sepsis before clinical diagnosis, up to 24 hours before the time of blood culture. External validation confirmed that these signatures may be generalized for sepsis detection at centers with variable patient populations, monitoring equipment, and practice patterns. Additionally, we found that static demographic variables help to risk-stratify infants at baseline but dampen the dynamic risk prediction.

### A cardiorespiratory signature of neonatal sepsis

Previously, we showed that a signature of sepsis exists in the HR signal^9,30^. Decreased HR variability with transient HR decelerations was recognized as similar to patterns of fetal distress that also served as a physiologic biomarker of sepsis in premature infants^31^. A decade of prior work translated abnormal HR patterns into mathematical algorithms ^7,8,32^, produced a validated predictive model for LOS in VLBW infants ^32^, known as the HRC index or HeRO score, and demonstrated reduced mortality in a multicenter randomized trial^1^. The past work exemplified a pathway for the translation of ideas about signatures of illness in continuous cardiorespiratory monitoring data into bedside tools for clinicians and patients.^33^

More recently, our group studied control of ventilation in premature infants and recognized that the HR and SpO_2_ signals correlate when episodes of apnea or periodic breathing lead to decreases in both HR (bradycardia) and SpO_2_ (desaturation) ^3,34^. Addition of the HR-SpO_2_ cross-correlation coefficient improved the performance of statistical models to detect sepsis over HRC monitoring alone in a two-NICU cohort. We found HR-SpO_2_ cross-correlation to be the best individual feature to discriminate LOS vs. sepsis-ruled out events.^5^ We also compared vital signs across our three collaborating sites and found clinically trivial though statistically significant differences in HR and SpO2^35^.

Respiratory deterioration prompts a majority of the sepsis evaluations in VLBW infants. Features in the POWS model quantify HR and SpO_2_ signals and their interaction, thereby capturing reduced variability, decelerations, desaturations, and other more subtle components of a signature of cardiorespiratory deterioration due to sepsis.

These findings support the idea that analysis of respiratory data, in addition to heart rate data, can enhance early detection of subacute but potentially catastrophic illnesses such as sepsis.

### Machine learning methods yield similar results

Interestingly, the choice of machine learning methods did not significantly impact performance. Logistic regression performed comparably to more complex methods. Unlike our prior work, ^3,5^ here we used cubic splines to account for non-linear relationships of predictors. We hypothesize that this resulted in similar performance of logistic regression compared with other machine learning methods that allow for non-linear relationships. Given equal performance but better explainability, we used the logistic regression approach for the remainder of the analysis. Our finding is matched by others in the literature; previous analyses of multiple machine learning methods for detecting LOS have also found similar performance across modeling techniques ^36,37^.

### Comparison with other work

Other groups have used machine learning methods and high-resolution cardiorespiratory data to develop models predicting LOS. Researchers in the Netherlands developed novel algorithms to detect reduced infant motion^38,39^ and demonstrated improved LOS prediction when combined with features measuring heart rate, SpO2, and respiratory rate^40^. We also find that combining multiple physiological features improves LOS prediction and extended the analysis to larger and external cohort. Other studies have included respiratory rate (RR)^39,41^, but our prior work has shown RR-derived features to be less predictive in sepsis models^3^.

Studies have repeatedly shown clinical variables and laboratory values to add to vital sign data for sepsis prediction^5,36,41–43^. We found that demographic variables improve model AUCs, but dampen the dynamic performance of a continuous sepsis risk model. Physiologic data is continuous, objective, and contains the earliest signs of the inflammatory response to infection via the autonomic nervous system^44^. While including demographic variables or clinician-initiated laboratory data may result in higher AUC, it may limit the lead time for early detection of sub-acute deterioration due to sepsis.

### Limitations

We captured a large number of infants and events, but some infants and events were excluded due to missing data. At NICU 2, the data were missing at random, while at NICU 3 the data retrieval process favored the smaller, sicker infants (less likely to change rooms, interrupting the flow of data). Prospective analysis with comprehensive data capture is needed to test models and risk trajectories over time and episodes in a multicenter cohort of premature infants.

Definitions of neonatal sepsis in premature infants vary widely among published studies^45,46^ due to challenges in developing a consensus definition for sepsis in this unique population^47^. We chose to model events of culture-proven sepsis, knowing that this definition, while unequivocal, also excludes many clinically important events associated with a negative blood culture. The heterogeneity of non-culture proven events likely impacts model development^41^ and, therefore, performance on new data. Nevertheless, our models trained exclusively on culture-positive events displayed a rise in predicted risk near the time of negative blood cultures diagnosed as clinical sepsis or NEC, indicating utility for detecting deterioration associated with sepsis-like events.

### Clinical implications and future directions

The goal of this work is to optimize a physiology-based sepsis early warning system to improve outcomes by bringing the clinician to the bedside of the right patient at the right time, even when resources are limited to pulse oximetry. All too often, LOS diagnosis occurs after the infection progresses to an advanced phase of systemic inflammation, organ dysfunction, and shock. Our results show that cardiorespiratory predictive monitoring can detect a sub-clinical prodrome in HR and SpO_2_ data with superior discrimination and sensitivity compared to analytics from either signal alone or demographic risk factors. External validation of POWS in two geographically distinct cohorts indicates that the signatures of sepsis may be general and are not greatly impacted by center-specific practice patterns or equipment. The ultimate test for validating our models and findings will be a prospective, multicenter clinical trial to measure the impact on clinical care and outcomes.

## Conclusion

A cardiorespiratory early warning score, analyzing heart rate from electrocardiogram or pulse oximetry together with SpO_2_, predicts late-onset sepsis diagnosis within 24h across multiple NICUs and detects sepsis better than heart rate characteristics or demographics alone.

## Data Availability

All data produced in the present study are available upon reasonable request to the authors

## Data Availability

The datasets generated during and/or analyzed during the current study are available from the corresponding author on reasonable request.

## Author Contributions

SK, BS, KF, DL, RS, ZV, and JRM have made substantial contributions to the conception or design of the work; SK, JQ, JB, AP, AB, and JI made substantial contributions to the acquisition, analysis, or interpretation of data; SK and BS drafted the work and all other authors have substantively revised it. All authors have approved the submitted version. All authors have agreed both to be personally accountable for the author’s own contributions and to ensure that questions related to the accuracy or integrity of any part of the work, even ones in which the author was not personally involved, are appropriately investigated, resolved, and the resolution documented in the literature.

## Consent Statement

This study was approved by the IRB at each site with waiver of consent.

