## Supplement for "Cardiorespiratory signature of neonatal sepsis: Development and validation of prediction models in 3 NICUs"

### Appendix

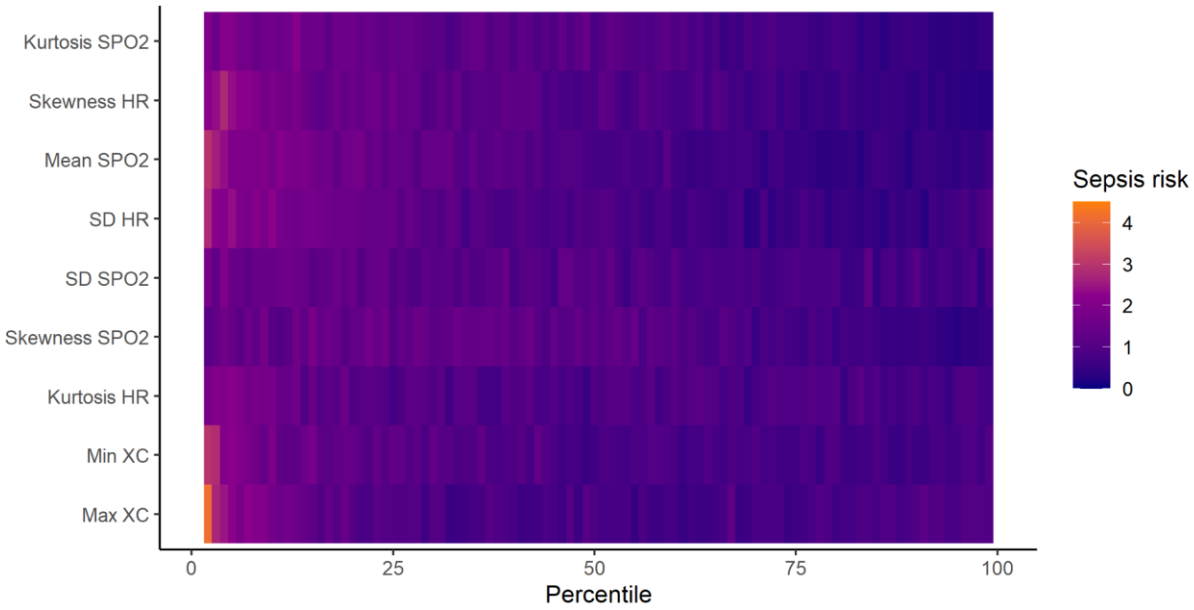


**S1 Fig: Candidate predictors and their empirical relationship with the outcome of sepsis at NICU 1**. Heat map depiction of the univariate risk of sepsis as a function of the nine HR and SPO2 features. Individual variables are on the y-axis and all observed values are displayed as deciles on the x-axis. Variables are in ascending or descending order based on highest risk. Orange color saturation indicates higher relative risk of the decile of data; a purple color indicates a lower relative risk.

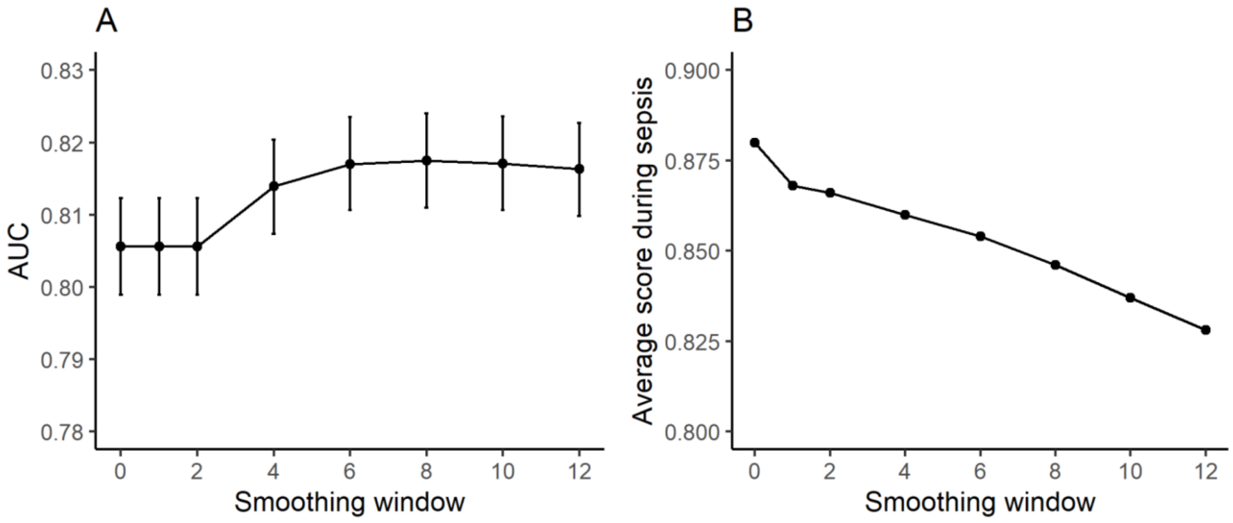


**S2 Fig. The effect of smoothing on model performance.** (A) displays the AUC as a function of different smoothing windows (in hours). (B) displays the average model output (presented as the z-score of the logit of the logistic regression model output) as a function of different smoothing windows.

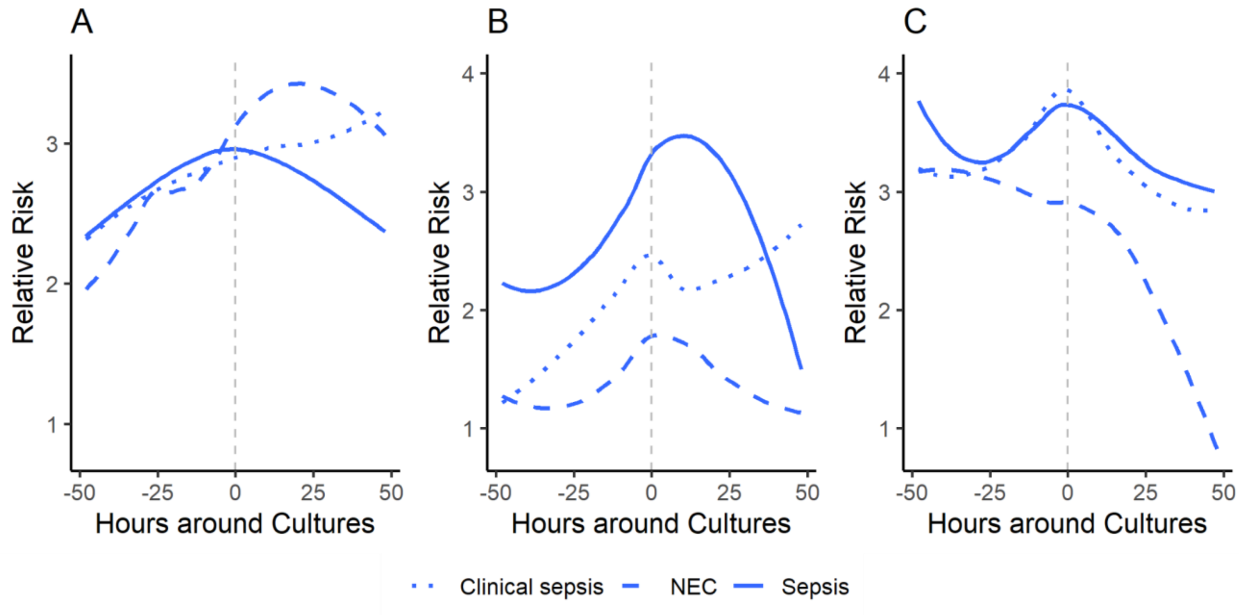


**S3 Fig. Average hourly POWS LR model relative risks around the time of blood cultures for clinical sepsis and NEC (without bacteremia).** Average hourly predictions for patients with clinical sepsis, sepsis, and NEC at (D) NICU 1, (E) NICU 2, and (F) NICU 3.

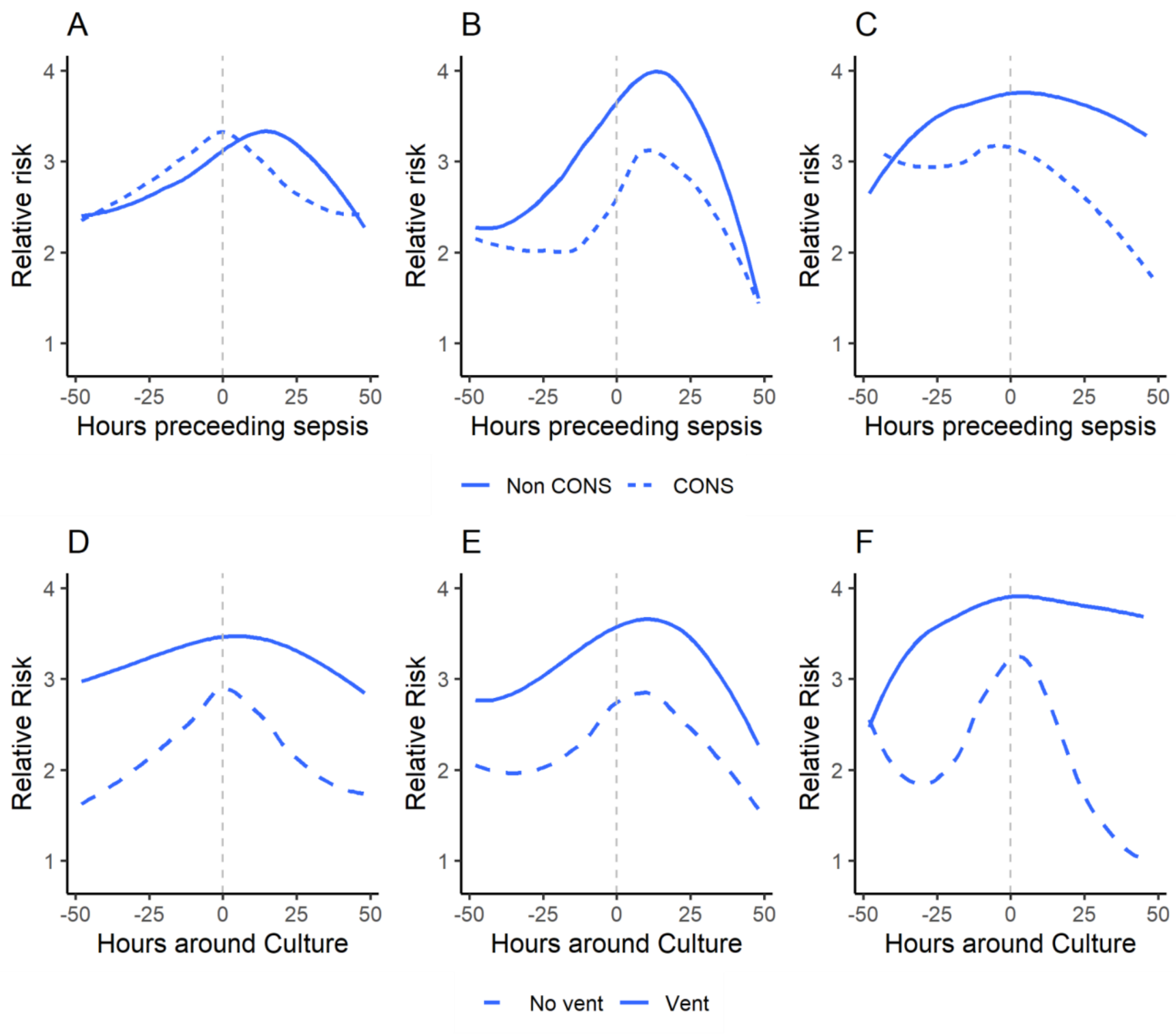


**S4 Fig. POWS LR model relative risks in CONS versus non-CONS and ventilator versus no ventilator.** Mean hourly POWS LR predictions are shown in the 48-hour period before and after the time of positive culture. From top left: average hourly predictions for CONS (dashed line) and non-CONS (solid line) at (A) NICU1, (B) NICU 2, and (C NICU 3. Average hourly predictions for patients ventilated at the time of positive blood cultures (solid line) and not ventilated (dashed line) for (D) NICU 1, (E) NICU 2, and (F) NICU 3.

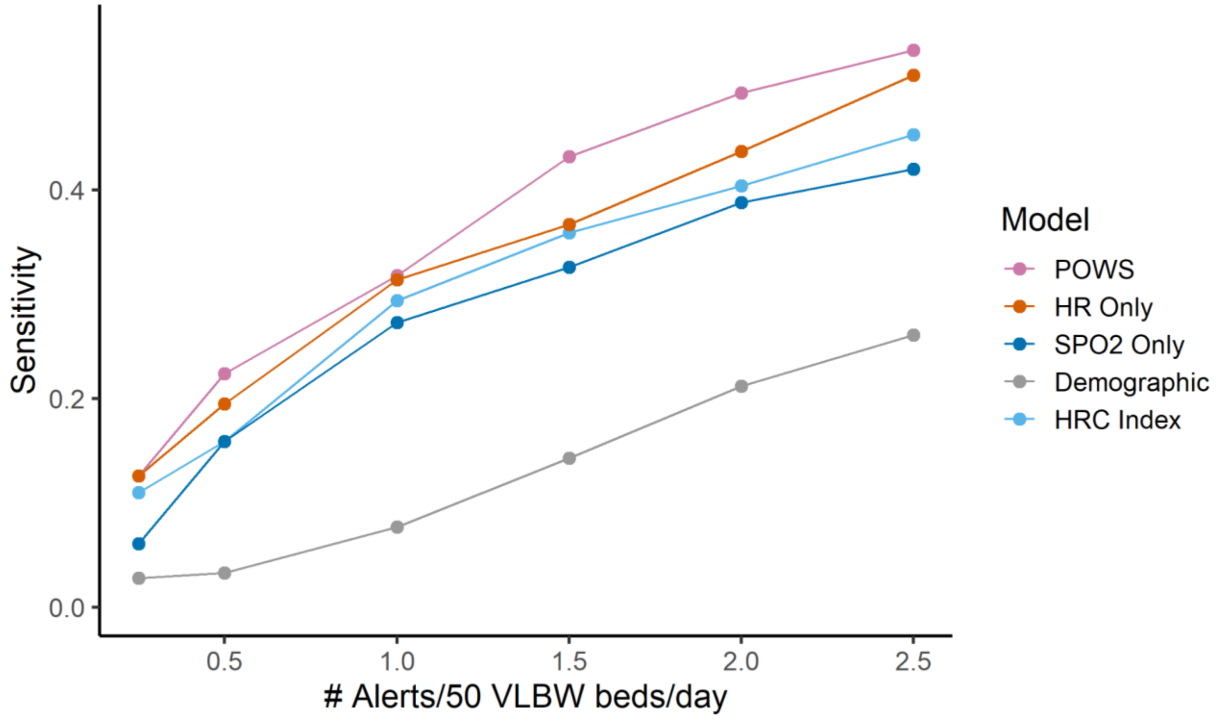


**S5 Fig. Evaluating the sensitivity of models across a range of thresholds.** We selected a range of thresholds and then calculated the number of alerts per day and required an alert to start within the day preceding the clinical diagnosis of sepsis. We defined alerts as daily threshold crossings. We excluded alerts in the seven days following a sepsis event. The y-axis displays the percent of sepsis events detected when allowing for different numbers of alerts per day. The HRC index performs similarly to the HR-only model despite differences in signal processing and features.

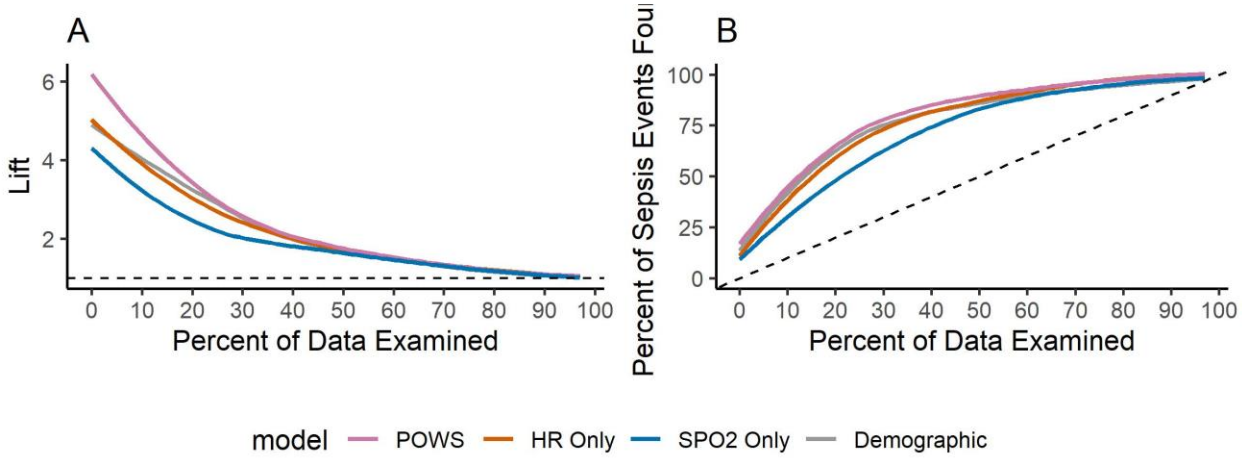


**Fig S6. Lift and Gain Charts**. Gain at a given decile is the ratio of cumulative events to the total number of events. Here, we can identify 67% of sepsis events by looking at 20% of the total data using both the HRC index and POWS models. The HR-only model identifies 57% of the events when looking at 20% of the total data. Lift is the ratio of gain % to random expectation % at a given decile. The figure can be interpreted as follows- a lift of 4 for the top decile of data means that when selecting 10% of data based on the model, you can expect 4x the total number of events found compared to randomly selecting 10% of data. You have a greater lift with the POWS models than the HR-only models when selecting 10% of data based on the model. For comparison, a lift of one is non-informative and a diagonal line on the gain chart is non-informative, represented by dashed lines.

Supplementary Tables

| Characteristic | log(OR) |
|---|---|
| Intercept | 1.115 |
| Mean $SpO_2$ | -0.057 |
| SD HR | -0.341 |
| SD HR (spline 1) | 0.354 |
| Skewness HR | -0.631 |
| Skewness HR (spline 1) | 0.218 |
| Skewness $SpO_2$ | 0.207 |
| Kurtosis HR | 0.053 |
| Kurtosis $SpO_2$ | -0.219 |
| Kurtosis $SpO_2$ (spline 1) | 0.453 |
| Max Cross Correlation | -0.994 |
| Max Cross Correlation (spline 1) | 1.873 |

**S1 Table. Logistic regression "POWS" model parameters.**
OR = Odds Ratio. Restricted cubic spline knot positions are: SD HR = 2.91, 6.27, 12.85; Skewness HR = -2.10, -0.03, 0.99; Kurtosis $SpO_2$ = 2.19, 3.94, 13.82; Max Cross-Correlation = -0.29, 0.07, 0.42

| Characteristic | log(OR) |
|---|---|
| Intercept | -2.551 |
| Birthweight (grams) | -0.002 |
| Age (days) | -0.048 |
| Age (spline 1) | 0.045 |

| | |
|---|---|
| Sex | |
| Female | -- |
| Male | 0.439 |

**S2 Table. Demographic model parameters.**
OR = Odds Ratio. Restricted cubic spline knot positions are: Age = 11, 46, 128

| Characteristic | log(OR) |
|---|---|
| Intercept | -4.353 |
| SD HR | -0.353 |
| SD HR (spline 1) | 0390 |
| Skewness HR | -0.996 |
| Skewness HR (spline 1) | 0.366 |
| Kurtosis HR | -0.144 |
| Kurtosis HR (spline 1) | 0.128 |

**S3 Table. Heart rate (HR) model parameters.**
OR = Odds Ratio, Restricted cubic spline knot positions are: SD HR = 2.91, 6.27, 12.85; Skewness HR = -2.10, -0.03, 0.99; Kurtosis HR = 2.09, 3.47, 11.94

| Characteristic | log(OR) |
|---|---|
| Intercept | 1.53 |
| Mean $SpO_2$ | -0.065 |
| Mean $SpO_2$ (spline 1) | -0.099 |
| SD $SpO_2$ | -0.201 |
| SD $SpO_2$ (spline 1) | 0.227 |
| Skewness $SpO_2$ | -0.32 |

| | |
|---|---|
| Kurtosis $SpO_2$ | -0.265 |
| Kurtosis $SpO_2$ (spline 1) | 0.546 |

**S4 Table. $SpO_2$ model parameters**
OR = Odds Ratio, Restricted cubic spline knot positions are: Mean $SpO_2$ = 88.67, 95.03, 99.21; SD $SpO_2$ = 0.68, 2.08, 5.89; Kurtosis $SpO_2$ = 2.19, 3.94, 13.82

| Characteristic | log(OR) |
|---|---|
| Intercept | 2.726 |
| HRC index | 0.577 |
| POWS logistic regression model | 1.132 |

**S5 Table. Coefficients of a bivariate model using the POWS logistic regression model and the HRC index model predictions as input features.** The higher log(OR) indicants greater weight in or importance in sepsis predictions.
OR = Odds Ratio.
